# Age moderates resting-state functional connectivity alterations in chronic low back pain

**DOI:** 10.64898/2026.11.26.26369098

**Authors:** Madeleine Hau, Laura Sirucek, Christian F. Beckmann, Petra Schweinhardt

**Affiliations:** Department of Chiropractic Medicine, Integrative Spinal Research Group, Balgrist University Hospital, University of Zurich, Zurich, Switzerland; Neuroscience Center Zurich, University of Zurich, Zurich, Switzerland; Center for Neuroplasticity and Pain (CNAP), Department of Health Science and Technology, Aalborg University, Aalborg, Denmark; Donders Institute for Brain, Cognition and Behaviour, Radboud University Medical Center, Nijmegen, The Netherlands; Oxford Centre for Functional Magnetic Resonance Imaging of the Brain (FMRIB), University of Oxford, Oxford, United Kingdom

## Abstract

Chronic low back pain (cLBP) is a complex musculoskeletal condition that often involves disrupted resting-state functional connectivity (rsFC). An important confounding factor to rsFC is age, as rsFC has been shown to decrease during healthy aging. Literature on grey matter volume (GMV) indicates accelerated brain aging in chronic pain patients compared to healthy controls (HC). However, it remains unclear whether such putative accelerated brain ageing in chronic pain is also reflected in rsFC. This study investigated differences in the effect of age on rsFC and GMV between cLBP patients and HC.

Forty-five cLBP patients and 34 age- and sex-matched HC underwent a 12-min whole-brain functional resting-state and a structural scan on a 3T MR-scanner. RsFC was analyzed using independent component analysis and dual regression, and GMV using voxel-based morphometry. An age by cohort interaction analysis on rsFC and GMV was performed using permutation tests.

A significant age-cohort interaction effect was detected on rsFC between attention, visual networks and brain regions in sensorimotor, parietal, frontal and cerebellar cortices. Across all significant clusters, younger patients showed increased rsFC, whereas older patients showed decreased rsFC compared to HC. There was no significant interaction nor group effect on GMV.

These findings indicate that differential age effects between cLBP patients and HC extend beyond structural measures and suggest an age-dependent change in attentional processing of sensorimotor stimuli rather than accelerated brain aging as potential mechanism for the observed changes. This demonstrates that age should be considered a variable of interest instead of a simple confounder.

**Significance:** Age is usually treated as a confounder in rsFC studies, corrected for across cohorts, despite structural data indicating differential age effects in cLBP patients compared to healthy controls. By demonstrating a significant age by cohort interaction effect on rsFC, the present study highlights the importance of investigating age effects within rather than across cohorts. Because mis-specified interactions can produce biased estimates of main effects and inflate error rates, age by cohort interactions should be examined to interpret rsFC findings accurately and draw sound conclusions about group differences. Further, investigating age by cohort interactions may help reveal potential age-dependent mechanisms in cLBP.

## Introduction

Chronic low back pain (cLBP), one of the leading causes of disability worldwide (Ferreira et al., 2023), is increasingly recognized as a musculoskeletal disorder with complex mechanisms that extend beyond peripheral nociceptive drivers and involve alterations in the central nervous system (CNS) (Sanzarello et al., 2016; Arendt-Nielsen et al., 2018; Sirucek et al., 2024).

An established method to investigate how chronic pain affects the brain as part of the CNS is resting-state functional connectivity (rsFC). RsFC is assessed by using functional magnetic resonance imaging (fMRI) to measure spontaneous fluctuations in brain activity. Brain regions with highly correlated spontaneous activity are considered being functionally connected and form resting-state networks that are associated with specific functions (Biswal et al., 1995; Beckmann et al., 2005). An extensive body of literature has investigated rsFC in cLBP, with more than 60 studies consistently reporting alterations across multiple resting-state networks (Baliki et al., 2012; Baliki et al., 2014; Mao et al., 2022a; Baumbach et al., 2022; Mao et al., 2023).

An important contributing factor to rsFC is the participant’s age (Biswal et al., 2010; Khan et al., 2025). It has been shown that rsFC of default mode (Hafkemeijer et al., 2012; Tomasi and Volkow, 2012), salience (Onoda et al., 2012), sensorimotor (Wu et al., 2007), visual (Yan et al., 2011) and other resting-state networks (Allen et al., 2011) decreases with healthy aging (Allen et al., 2011; Ferreira and Busatto, 2013). While most studies report age-related decreases, a few also showed age-related increases, for instance in frontal, parietal and sensorimotor brain regions (Biswal et al., 2010; Tomasi and Volkow, 2012; Toussaint et al., 2014). Therefore, age is often considered a confound in neuroimaging studies and is usually treated as a nuisance regressor during the analysis (Kregel et al., 2015; Veinot et al., 2025; Sunavsky et al., 2025). This approach implicitly assumes that age effects are consistent across cohorts, e.g. patients and controls. However, a growing body of literature on *structural* brain measures indicates that age effects might differ between patients with chronic pain and healthy controls. Specifically, studies on grey matter volume (GMV) reported larger age-related decreases in patients with fibromyalgia (Kuchinad et al., 2007; Ceko et al., 2013), migraine (Chong et al., 2014), temporomandibular disorder (Moayedi et al., 2012) and cLBP (Yu et al., 2021) compared to healthy controls, often interpreted as accelerated brain aging. Thus, it is conceivable that age effects on other neuroimaging measures might also differ between patient cohorts and healthy controls. To the best of our knowledge, this has not been formally tested for rsFC in cLBP. The present study investigated whether the effects of age on rsFC differ between patients with cLBP and healthy controls as a secondary analysis within a larger project. Patients with cLBP and age- and sex-matched healthy controls underwent an MRI session that included whole brain structural and functional resting-state scans. Age by cohort interaction analyses were performed on the rsFC data as well as on the GMV in order to assess if potential functional changes are accompanied by structural changes.

## Methods

### 1. General

The present study is a secondary analysis conducted as part of a larger project within the Clinical Research Priority Program (CRPP) Pain (https://www.crpp-pain.uzh.ch/en.html). The CRPP Pain project investigated pain phenotypes across different chronic pain conditions in comparison to healthy participants. During three visits, lasting approximately 3h each, participants underwent psychophysical, neurophysiological, and MR assessments.

The study was approved by the local ethics committee and was performed according to the guidelines of the Helsinki Declaration (EK-04/2006/PB_2016-02051 and PB_2019-00136, clinicaltrial.gov number: NCT02138344 and NCT04433299). All participants gave their written informed consent before participating.

### 2. Study Overview

Within the larger study, participants completed two MR sessions: one comprising a functional resting-state and structural data acquisition (resting-state session), the other a MR spectroscopy (MRS) and structural data acquisition (MRS session). This study used the functional data from the resting-state session and the structural data from the MRS session. The latter was done because the image contrast of the structural scan of the MRS session was better compared to the structural data of the resting-state session. The MRS session was conducted approximately 2 hours before or after the resting-state session for all but three participants, where it was conducted on a separate day (3, 7, and 28 days later, respectively). For 3 subjects, who did not participate in the MRS session, the structural data from the resting-state session was used.

Immediately after leaving the scanner, participants were asked if they experienced any pain (including low back pain for the patients) throughout the scan on a 11-point numerical rating scale (NRS) ranging from 0 (“not painful”), 1 (“just painful”) to 10 (“most intense pain tolerable”).

### 3. Participants

CLBP patients and healthy controls aged between 18 and 80 were included in the present analysis. Details on the recruitment process and the in- and exclusion criteria have been published previously in (Sirucek et al., 2024). In brief, participants were recruited via the department’s policlinic, online advertisement and word of mouth. All cLBP patients needed to have low back pain for at least three months with cLBP being the primary source of pain and without any signs of underlying serious pathology or radiculopathy. Healthy controls may not have had low back pain on more than 3 consecutive days in the last year. Exclusion criteria were any chronic pain condition (except cLBP for the patients), any major medical or psychiatric condition, pregnancy or inability to follow study instructions. All participants were asked to report demographic information and underwent a clinical examination. CLBP patients reported the average pain intensity of their back pain during the last four weeks on the same 11-point NRS as described above (see Study Overview) and the duration of their pain in months.

### 4. Neuroimaging

### 4.1. Data acquisition

The resting-state session was performed on a 3 Tesla Siemens MAGNETOM Prisma scanner using a 32-channel head coil (Siemens Healthcare, Forchheim, Germany).

The session included a structural scan, a field map using a gradient echo sequence (TR: 649.0 ms, TE: 4.92 ms /7.38 ms, voxel size: 2.2 mm isotropic, field of view: 210 mm) and a 12 min whole-brain functional resting-state scan using a T2*-weighted echo-planar imaging (EPI) sequence (repetition time (TR): 846 ms, echo time (TE): 30 ms, voxel size: 2.2 mm isotropic, field of view: 210 mm, flip angle: 56°, 840 volumes, multiband acceleration factor: 6). As described above, the structural scan was not used for most of the subjects (for sequence parameters and subject information see Supplementary material, Methods S1). Participants were instructed to lie still in the scanner with eyes open focusing on a fixation cross, while letting their mind wander (i.e., without focusing on any specific thoughts or performing cognitive demanding tasks such as mental arithmetic). The total session lasted approximately 20 min.

The MRS session was performed on a 3 Tesla Philips Achieva scanner using a 32-channel head coil (Philips Healthcare, Best, The Netherlands). The session included a 7 min T1-weighted structural scan using a turbo field echo (TFE) sequence (TR: 8.1 ms, TE: 3.7ms, inversion time (TI): 1014.77ms, voxel size: 1 mm isotropic, FOV: 240 mm, flip angle: 8°, number of TFE shots: 150; TFE shot duration: 1999.9 ms) and an MRS scan, which is not part of the present analysis. Where available, this structural data was used for the present analysis.

#### 4.2. Preprocessing and Analysis

Functional and structural data was preprocessed and analyzed using FSL version 6.0.4 (Jenkinson et al., 2012).

##### 4.2.1. Functional Data

The preprocessing was conducted using FSL FEAT (Woolrich et al., 2001). It included motion correction with FSL MCFLIRT (Jenkinson et al., 2002) and a high-pass temporal filter with a cut-off of 2000 s to remove slow signal drifts. Next, denoising was conducted on the minimally preprocessed functional data by performing an unrestricted Independent Component Analysis (ICA) on the single subject data using FSL melodic (Beckmann and Smith, 2004). Subsequently, the obtained independent components (ICs) were classified as “signal”, “noise” or “unknown” using the supervised machine learning classifier FSL FIX (Salimi-Khorshidi et al., 2014), which was pre-trained on the Human Connectome dataset (HCP25_hp2000 training file; threshold = 20) (Smith et al., 2013). A random sample of the classifications was reviewed for accuracy. Components classified as “noise” were regressed out.

The denoised functional data was registered to the structural image obtained on the Philips scanner during the MRS session. For this, the structural image was first registered to the structural image obtained during the resting-state session with a rigid body transformation and the default correlation ratio cost function using FLIRT (Jenkinson and Smith, 2001; Jenkinson et al., 2002). The functional data was registered to the structural image using a rigid body transformation with a boundary-based registration (BBR) cost function, which incorporated a correction for spatial distortions using field maps, and, subsequently, non-linearly transformed to MNI-152 standard space. Finally, smoothing with a 4 mm full-width-at-half-maximum (FWHM) Gaussian kernel was conducted to improve signal-to-noise ratio using the FSL utility tool fslmaths.

On the group level, an ICA restricted to 30 ICs was used to construct group-level resting-state networks and dual regression with variance normalization was performed to retrieve the single subject networks (Beckmann et al., 2005). Obtained ICs were visually inspected for correspondence with canonical resting-state networks (Beckmann et al., 2005; Seeley et al., 2007; Smith et al., 2009). Components representing group level noise (e.g., signal in white matter (WM) or cerebrospinal fluid (CSF)) were excluded from further analysis. After this, differences in age effects on rsFC between cLBP patients and healthy controls were assessed using an age by cohort interaction design. Both contrasts (slope of age in cLBP patients > slope of age in healthy controls, and vice versa) were tested using non-parametric permutation tests (5000 permutations) using FSL randomise (Winkler et al., 2014). Multiple comparison correction was performed using threshold-free-cluster-enhanced (TFCE, (Smith and Nichols, 2009)) family-wise-error (FWE) rate. Parameter estimates (PEs) of significant clusters were extracted as β-values for follow-up analysis and visualizations.

##### 4.2.2. Structural data

The structural data was analyzed according to the standardized FSL VBM pipeline (Ashburner and Friston, 2000; Good et al., 2001). In brief, brain extraction was performed, followed by segmentation into GM, WM and CSF. Subsequently, a study-specific GM template was created based on the structural data of all healthy controls and the same number of randomly selected cLBP patients and affine registered to standard space. Participants with structural data obtained during the resting-state session (N=3) were not used for the GM template. Next, the individual GM images were registered to the template and Jacobian modulation was used to compensate for the volume distortions introduced during the non-linear transformation. Finally, smoothing with a 3 mm FWHM Gaussian kernel was applied.

On the group level, an age by cohort interaction analysis, as described above, was performed to assess differences in age effects on GMV between cLBP patients and healthy controls. In case there was no significant interaction effect present, a group comparison corrected for age and sex was performed. To ensure a comparable sample in the functional and the structural analysis, the three participants with structural data from the resting-state session were included in all VBM analyses and *scanner type* (Siemens vs Philips) was used as an additional covariate. To examine if the three participants drove the statistical inference, the analysis was repeated without them. If exclusion changed the statistical inference, the results without these three participants are reported.

All analyses were conducted on the whole brain level, as well as with a region of interest (ROI) approach, in order to assess if age by cohort interaction effects on rsFC can be explained by age by cohort interaction effects on GMV. For the ROI approach, significant clusters from the rsFC analysis were binarized and combined to one mask for the VBM analysis. For both approaches, non-parametric permutation tests (5000 permutations) using FSL randomise and TFCE-enhanced FWE rate correction for multiple comparisons were implemented.

### 5. Further Statistical Analysis

Statistical analysis of demographic data and follow-up analysis on extracted PEs was conducted in R version 4.5 for Windows. A p-value < 0.05 was considered statistically significant. Normality of raw data or model residuals (depending on the used statistical test) was assessed by visual inspection of histograms and quantile-quantile (QQ)-Plots. Differences between cohorts in demographic data and pain experienced during the scan were assessed using t-tests for parametric data, Wilcoxon rank-sum tests for non-parametric data (i.e., data obtained using the ordinal NRS), and Chi-Square test for categorical data (i.e., sex).

As age and pain duration are often inherently linked, it was investigated if age by cohort interaction effects were influenced by pain duration. For this, it was first assessed if pain duration and age in cLBP patients were associated using a Pearson correlation for parametric, or a Spearman correlation for non-parametric data. If age and pain duration were correlated, it was assessed whether pain duration explained unique variance in rsFC or GMV beyond the effects of age in cLBP patients. For this, linear models using the lm() function for parametric data and rank-based models using the rfit() function of the Rfit package for non-parametric data, were performed to assess the effect of pain duration and age as covariate on the extracted PEs of significant clusters. Of note, the effect of age in these models was not interpreted because it can be inflated due to the nonindependence error (Vul et al., 2009). Additionally, because head motion is often associated with age, potential motion-related bias was assessed by comparing the effect of age on mean framewise displacement between groups using linear or rank-based models as described above.

## Results

### 1. Participants

Out of 64 cLBP patients and 39 healthy controls recruited for the larger project, 16 cLBP patients and 4 controls did not want to participate in the MRI assessment. Additionally, three patients were excluded due to non-compliance, and one healthy control was excluded due to abnormal sensory findings over the course of the larger project. Thus, the final sample analyzed here comprised 45 (25 female) cLBP patients and 34 (19 female) healthy controls. Demographic data, clinical characteristics, and pain experienced during the scan are reported in Table 1. Age distributions for each cohort are reported in the supplementary material (Figure S1).

**Table 1:** Participant Information. For parametric data, values are presented as mean, standard deviation (SD), and T and p values from two sample T-tests. For non-parametric data, values are presented as median, interquartile range, and W and p values from Wilcoxon rank-sum test, or absolute numbers, percentages, and χ^2^ and p values from chi-square tests for categorical data. Average pain intensity for the patients refers to the last four weeks. BMI, Body Mass Index; cLBP, chronic low back pain; NRS, numerical rating scale

|  | cLBP | Healthy controls | Test Statistic |
|---|---|---|---|
| Number of participants | 45 | 34 |  |
| Age [years]; mean, SD | 52.7, 16.73 | 50.5, 17.11 | T (70.4) = 0.58, p = 0.57 |
| Sex; number, % females | 25, 55.56% | 19, 42.22% | χ² < 0.01, p = 1 |
| BMI [kg/m<sup>2</sup>]; mean, SD | 20.8, 3.09 | 20.0, 2.45 | T (76.8) = 1.22, p = 0.23 |
| Pain during the scan (NRS); median, interquartile range | 0, 4 | 0, 0 | W = 1097.5, p < 0.01 |
| Low back pain characteristics |  |  |  |
| Average pain intensity (NRS); median, interquartile range | 4, 2 |  |  |
| Pain duration [months]; median, interquartile range | 106.0, 188.25 |  |  |

### 2. Age effects in attention, visual and auditory networks are consistently different in cLBP patients

Out of 30 ICs, 22 aligned with previously reported resting-state networks and were used for further analysis. Eight ICs were judged to be group level noise and were excluded from the analysis. Rank-based models showed that age effects on mean framewise displacement did not differ between cohorts and that mean framewise displacement was not associated with age or cohort, as indicated by non-significant interaction and main effects (interaction: b < 0.01, t = 0.64, p = 0.525; age: b < 0.01, t = -0.64, p = 0.523; cohort: b = -0.02, t = -0.67, p = 0.502).

A significant age by cohort interaction effect was detected in 6 ICs pertaining to 5 resting-state networks (p’s < 0.05, TFCE enhanced FWE correction), indicating that in these networks, the effect of age on rsFC was significantly lower in cLBP patients compared to healthy controls (slope of age in cLBP < slope of age in HC; Figure 1A). Descriptive analysis of the extracted PEs revealed that younger patients showed increased rsFC, whereas older patients showed decreased rsFC compared to age-matched healthy controls across all significant brain regions (Figure 1A; age distribution per cohort: Figure S1). No brain region showed a significant interaction effect in the opposite contrast (slope of age in cLBP > slope of age in healthy controls).

**Figure 1:**
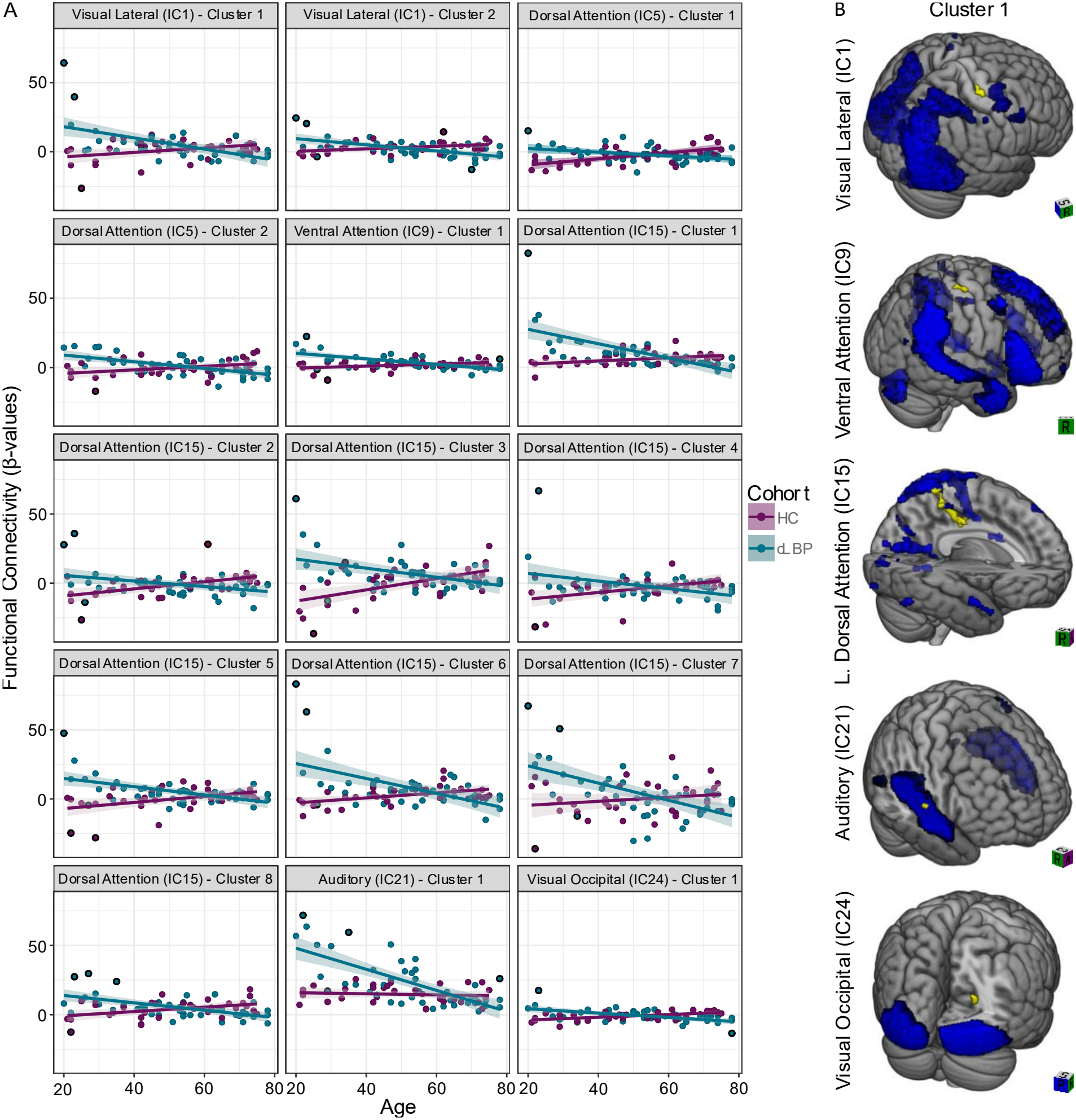
Age effects in attention, visual and auditory networks are consistently different in cLBP patients. **A:** Parameter estimates extracted from clusters with a significant age-by-cohort interaction effect, plotted against age for cLBP patients and healthy controls, raw values (dots), with fitted mean within cohort (solid lines) and 95% confidence intervals (shaded areas). Because the clusters were selected based on the significant interaction effect, the depicted slopes necessarily differ between cohorts and illustrate the direction of the effect. Data points with a strong influence on the illustrated effects were determined based on standardized residuals and Cook’s distance and are marked with a black outline. **B:** Brain regions showing age-related differences in rsFC with independent components between cohorts, yellow color indicates significant clusters (p < 0.05, TFCE-FWE corrected), blue color indicates group-level IC maps. Depicted are the largest cluster (cluster 1) and one IC map for each affected resting-state network. A complete list of all significant clusters is provided in Table 2. cLBP, chronic low back pain; HC, healthy controls; IC, independent component; L, left.

Significant age by cohort interaction effects were observed between the ventral attention (IC 9), left dorsal attention (IC 5 & 15), visual-lateral (IC 1), visual-occipital (IC 24) networks and several cortical and cerebellar brain regions (Figure 1B). These brain regions were located in primary somatosensory cortex (S1)/primary motor cortex (M1), superior and inferior parietal cortex, frontal pole, posterior cingulate cortex, visual cortex, and left crus of the cerebellum. In addition, a significant interaction effect was observed within the auditory network (IC 21) (Figure 1B). Detailed information on all significant clusters and related networks is provided in Table 2.

**Table 2:** Brain regions exhibiting a significant age by cohort interaction effect on rsFC in cLBP patients and healthy controls. Clusters are listed under the respective resting-state network in which the interaction effect was observed. For each cluster, peak t-values, TFCE enhanced FWE corrected p-values, peak coordinates in MNI space, and cluster size in number of voxels is reported. Anatomical labels were assigned using the FSL atlasquery function. Cyto- and myeloarchitectonic labels based on the Jülich Histological Atlas are provided where available. BA, Brodmann area; L, left; R, right

| Cluster | Brain Region | Side | MNI Coordinates |  |  | Cluster Size | Maximum T-value | P-value |
|---|---|---|---|---|---|---|---|---|
|  |  |  | x | y | z |  |  |  |
| Visual Lateral Network (IC 01) |  |  |  |  |  |  |  |  |
| 1 | Premotor cortex (BA6), S1 (BA1, BA3b, BA2), M1 (BA4a, BA4p) | R | 42 | -26 | 68 | 27 | 4.73 | 0.016 |
| 2 | Premotor cortex (BA6), M1 (BA4a, BA4p) | R | 32 | -22 | 68 | 9 | 4.32 | 0.035 |
| Left Dorsal Attention Network (IC 05) |  |  |  |  |  |  |  |  |
| 1 | Inferior parietal lobule (PGp) | R | 36 | -72 | 34 | 10 | 4.73 | 0.027 |
| 2 | Anterior intra-parietal sulcus (Area hIP1) | R | 36 | -64 | 34 | 6 | 4.45 | 0.039 |
| Ventral attention Network (IC 09) |  |  |  |  |  |  |  |  |
| 1 | Premotor cortex, M1 (BA6), S1 (BA1) | R | 42 | -18 | 62 | 29 | 5.01 | 0.015 |
| Left Dorsal Attention Network (IC15) |  |  |  |  |  |  |  |  |
| 1 | Superior parietal lobule (Area 5Ci, Area 5M, Area 5L, Area 7A), S1 (BA3a, BA3b), M1(BA4a, BA4p) | L | -12 | -44 | 50 | 160 | 4.72 | 0.008 |
| 2 | M1 (BA4a, BA4p), S1 (BA1, BA3a, b), Premotor cortex (BA6) | R | 34 | -24 | 54 | 38 | 3.9 | 0.029 |
| 3 | Inferior parietal lobule (Area PGp, Area PGa) | L | -50 | -72 | 20 | 32 | 4.81 | 0.012 |
| 4 | M1 (BA4p, BA4a), S1 (BA1, BA2, BA3b), Premotor cortex (BA6) | R | 42 | -16 | 54 | 25 | 3.63 | 0.038 |
| 5 | Visual cortex V2; V3V; V4 | R | 40 | -90 | 6 | 20 | 4.32 | 0.037 |
| 6 | Frontal Pole | R | 28 | 56 | 14 | 16 | 4.46 | 0.035 |
| 7 | Posterior cingulate gyrus | L | -8 | -20 | 28 | 7 | 3.82 | 0.042 |
| 8 | Cerebellar Crus I; Crus II | L | -22 | -86 | -30 | 5 | 4.8 | 0.035 |
| Auditory Network (IC 21) |  |  |  |  |  |  |  |  |
| 1 | Primary auditory cortex (TE1.0, TE1.2); S2/parietal operculum (OP1, OP4) | R | 60 | -8 | 4 | 11 | 4.89 | 0.023 |
| Visual Occipital Network (IC 24) |  |  |  |  |  |  |  |  |
| 1 | Visual cortex V1 (BA17); V2 (BA18) | R | 22 | -60 | 16 | 23 | 4.81 | 0.014 |

Spearman’s correlation analysis showed a significant positive correlation of age and pain duration in cLBP patients (rho = 0.37, p = 0.012), indicating that older patients had pain for a longer duration. Rank-based models revealed that there was no significant main effect of pain duration on the PEs of all significant clusters (Table S1), indicating that there was no evidence that pain duration explained variance in rsFC of cLBP patients beyond age.

### 3. No age-related or cohort difference in GMV between cLBP patients and healthy controls

There was no significant age by cohort interaction effect on GMV neither in the whole brain approach nor in the ROI approach (p > 0.05, TFCE enhanced FWE correction, Figure 2A), indicating that the effect of age on GMV did not differ between cLBP patients and healthy controls across the whole brain nor in brains regions exhibiting age-related rsFC changes. In addition, there was no significant difference in GMV between cLBP patients and healthy controls, across the whole brain or in the ROIs (Figure 2B). Exclusion of the three participants whose structural scan was acquired on a different scanner did not change the statistical inference, thus the results of the full sample are reported.

**Figure 2:**
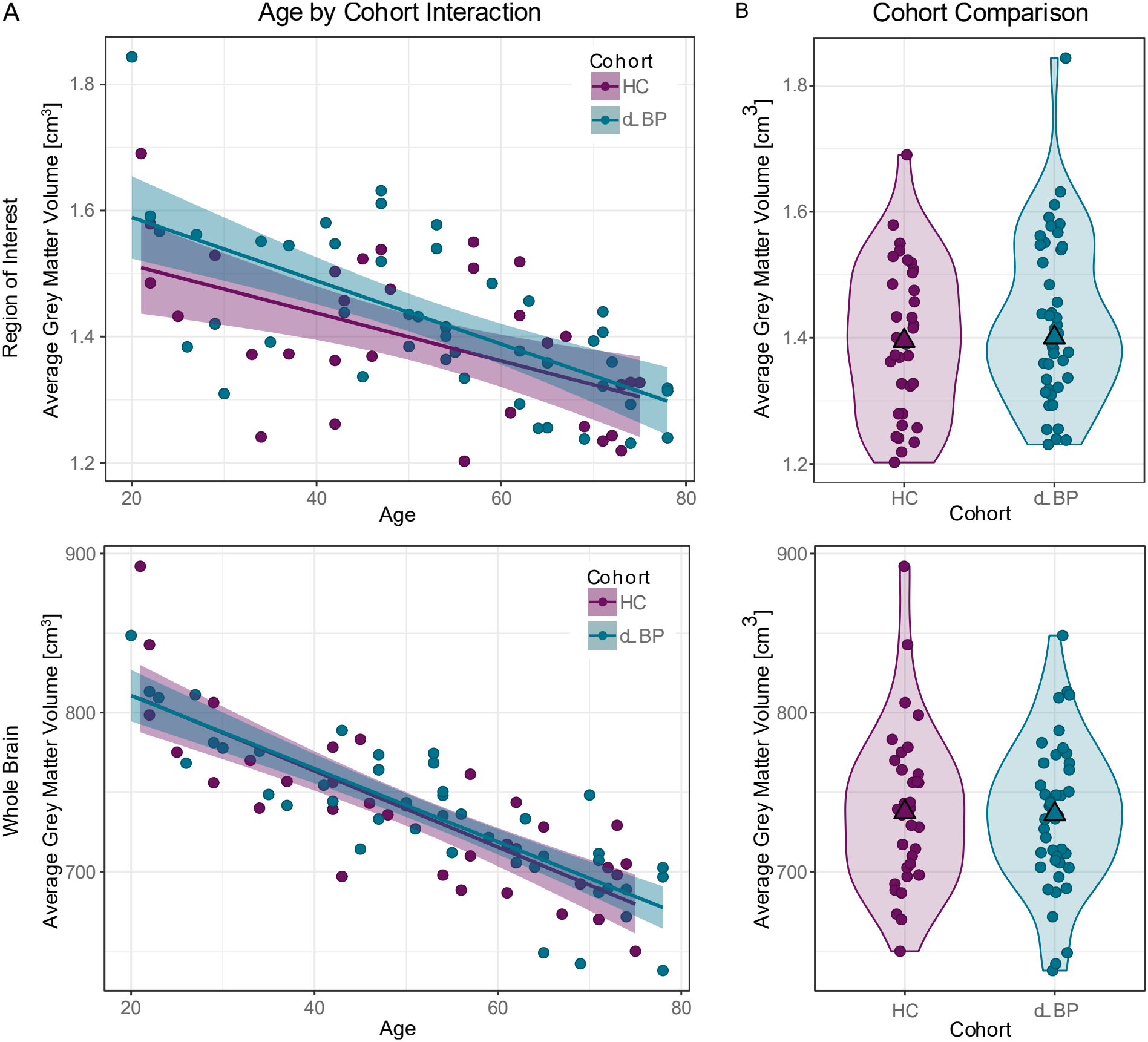
No age-related or cohort difference in GMV between cLBP patients and healthy controls. **A:** Average GMV plotted against age for cLBP patients and healthy controls in ROI consisting of all significant clusters from the rsFC analysis combined (upper panel) and across the whole brain (lower panel), with average GMV (dots), fitted regression line per group (solid lines) and 95% confidence interval (shaded areas). **B:** Cohort comparison corrected for age and sex in ROI (upper panel) and across the whole brain (lower panel), with average GMV (dots), median across cohorts (triangle), and density distribution of GVM for each cohort (shaded areas). cLBP, chronic low back pain; GMV, grey matter volume; HC, healthy controls

## Discussion

The present study aimed to investigate (1) if the effect of age on rsFC differs between cLBP patients and healthy controls by assessing age by cohort interaction effects on rsFC and (2) whether observed effects were related to GMV differences between cohorts. Significant age by cohort interaction effects were detected across several networks with patients showing larger age-related decreases of rsFC compared to healthy controls. Importantly, pain duration explained no unique variance in rsFC in cLBP patients and therefore likely did not confound the observed age by cohort interaction effects. In contrast, neither an interaction effect nor a cohort difference was detected for GMV.

The observed age by cohort interaction effect on rsFC extends prior studies on structural measures showing differential age effects on GMV across several chronic pain conditions compared to healthy controls (Kuchinad et al., 2007; Moayedi et al., 2012; Ceko et al., 2013; Chong et al., 2014; Cruz-Almeida et al., 2019; Yu et al., 2021; Lee et al., 2026). Also in line with the present results, a previous study on functional data investigated rsFC across cLBP patients of different age groups, without a comparison to healthy controls, and reported reduced rsFC in older compared to younger patients (Zu et al., 2024). Furthermore, amplitude of low-frequency fluctuation (ALFF) patterns used to classify cLBP have been shown to differ between younger and older patients (Baran et al., 2022). Together, these findings support the notion that differential age effects in cLBP compared to healthy controls are not restricted to structural measures but extend to functional measures, with important methodological implications for rsFC analyses. Previous studies focusing on group differences between patients and healthy controls included age as a nuisance regressor and/or included an age-matched control group to control for age effects, which implicitly assumes constant age effects across groups. While the vast majority of these studies report that group differences in rsFC between patients and healthy controls exist, there is large variability regarding the directionality of the change (Lee et al., 2026), with some studies reporting increased (Biswal et al., 2010; Tagliazucchi et al., 2010; Loggia et al., 2013; Yu et al., 2014; Kim et al., 2019; Letzen et al., 2020; Mao et al., 2022a; Mao et al., 2022b; Mao et al., 2023; Fan et al., 2023; Zhang et al., 2023; Zhu et al., 2024; Li et al., 2025b; Zhang et al., 2025), others reporting decreased rsFC in cLBP (Balenzuela et al., 2010; Yu et al., 2020; Ng et al., 2021; Ayoub et al., 2021; Baumbach et al., 2022; Li et al., 2025a; Zhao et al., 2025; Gu et al., 2025). This variability might partly be due to modelling age as a constant effect, because misspecification of interactions can lead to both attenuated or reversed effects, as well as inflated Type I error rates (Rimpler et al., 2025).

For GMV, larger age-related decreases are sometimes interpreted as accelerated brain aging (Kuchinad et al., 2007; Moayedi et al., 2012). Although GMV and rsFC can vary independently (Mowinckel et al., 2012; Schulz et al., 2022), several studies reported that they exhibit shared spatial patterns (Smith et al., 2019), network-specific correlations (Segall et al., 2012; Vidal-Piñeiro et al., 2014; Liu et al., 2022) and concurrent declines during healthy aging (Du et al., 2025). Therefore, it may be plausible to assume similar mechanisms underlying age-related differences between chronic pain patients and healthy controls in GMV and rsFC. However, the observed shape of the interaction (increased rsFC in younger and decreased in older patients compared to healthy controls), does not support this conclusion. Rather, a stronger slope in the same direction as in healthy controls, would have been expected for the effect of age on rsFC in cLBP patients.

In addition, the involved networks also suggest a different explanation for the findings of the present study. Differential age effects on rsFC were predominantly observed between the dorsal attention, ventral attention, visual networks and brain regions in S1/M1 and the inferior parietal cortex, as well as within the auditory network. These brain regions may indicate a shift from increased to reduced attentional processing of sensorimotor signals in cLBP with ageing. In general, chronic pain is associated with increased attentional processing of somatosensory signals, as suggested by the selective attention towards somatosensory stimuli, a phenomenon termed attentional bias (Roelofs et al., 2002; Todd et al., 2018; Broadbent et al., 2021; Robles et al., 2024). With age, this attentional bias may decline. In addition to functional data, this idea is supported by the post hoc finding that patients’ attention to their back pain (rated according to Akin-Akinyosoye et al., 2018) was negatively associated with age in the present study (Figure S2). This interpretation is supported by age-related decreases in body awareness (Ciaramella et al., 2023), pain catastrophizing (Murray et al., 2021), kinesiophobia (Cook et al., 2006), and increases in pain acceptance and self-efficacy observed in various chronic pain conditions (Sielski et al., 2017; Murray et al., 2021). Thus, attentional bias might be age-dependent with the observed connectivity changes in attention-related networks representing a possible neurophysiological mechanism underlying this phenomenon.

In contrast to previous findings, neither an interaction effect nor a cohort difference in GMV was observed. One potential explanation is that the patients in the present study represented a relatively high-functioning cLBP sample, characterized by moderate pain levels, high physical activity, absence of mood disorders, and low disability scores. In fact, Johnsen and colleagues reported that differences in brain-predicted age, an imaging biomarker of brain aging derived from GMV, were only significant in patients with high-but not low-impact knee pain (Johnson et al., 2022). The absence of GMV evidence for accelerated brain aging in the present sample might indicate that the differential age effects on rsFC might not reflect accelerated brain aging, but an age dependency in CNS processes associated with cLBP.

### Limitations

The present study was not primarily designed to assess age by cohort interaction effects but represents a secondary analysis within a larger study. Due to the exploratory nature and the absence of an a priori hypothesis regarding which resting-state networks might exhibit differential age effects, analyses were performed across all components associated with resting-state networks. For this exploratory setting, applying a Bonferroni correction across the components of the ICA was not considered feasible, because this highly conservative correction would likely obscure potential meaningful effects. Thus, the observed results require replication in studies specifically designed and powered to test age by cohort effects.

### Conclusion

In sum, the present study showed that the effect of age on rsFC differs between patients with cLBP and healthy controls and should therefore be investigated or controlled for within, rather than across groups in neuroimaging studies. The observed interaction shape and the networks involved might suggest an age-dependent shift in attentional processing in cLBP, rather than accelerated brain aging. Future studies could explore if these findings have implications for the effectiveness of cognitive-behavioral or attention-focused interventions across different age ranges. Finally, the present results highlight the importance of treating age not solely as a confounding variable but as a factor of interest, in order to better explain variability between subjects, avoid misinterpretation of main effects in statistical modelling, and improve our understanding of potential age-dependent mechanisms of cLBP.

## Data Availability

All data produced in the present study are available upon reasonable request to the authors

## Acknowledgment

This study was funded by the Clinical Research Priority Program ‘PAIN’ of the University of Zurich. L.S. was supported by the Theodor und Ida Herzog-Egli Stiftung and is affiliated with the Center for Neuroplasticity and Pain (CNAP), inaugurated by the Danish National Research Foundation (DNRF121) and is supported by a Swiss National Science Foundation Postdoc.Mobility Fellowship (P500-3_235289/1). We thank all participants who took part in the study. Additionally, we thank Balgrist Campus and the Swiss Center for Musculoskeletal Imaging for their support and providing the infrastructure to perform the present study.

