## Supplementary Material for "Age moderates resting-state functional connectivity alterations in chronic low back pain"

**MethodsS1: Sequence parameters of structural image acquired on the Siemens scanner**

The resting-state session included a 4.5 min T1-weighted scan using magnetization prepared rapid acquisition gradient echo (MP2RAGE) sequence (repetition time: 2300 ms, echo time: 2.25 ms, inversion time: 900 ms, voxel size: 1 mm isotropic, field of view: 240 mm, flip angle: 8°) on a 3 Tesla Siemens MAGNETOM Prisma scanner. The structural data acquired during this scan was used for three out of 79 participants (2 cLBP patients and 1 healthy control).

| Cluster | Effect of Pain duration | | | Effect of Age | | |
| --- | --- | --- | --- | --- | --- | --- |
|  | Estimate | T-value | p-value | Estimate | T-value | p-value |
| Visual Lateral Network (IC 01) | | | | | | |
| 1 | < 0.01 | 0.07 | 0.947 | -0.23 | -3.52 | 0.001 |
| 2 | - 0.01 | -1.66 | 0.105 | -0.14 | -2.94 | 0.005 |
| Left Dorsal Attention Network (IC 05) | | | | | | |
| 1 | < -0.01 | -0.23 | 0.818 | -0.12 | -2.34 | 0.024 |
| 2 | < 0.01 | 0.93 | 0.358 | -0.25 | -5.38 | < 0.001 |
| Ventral attention Network (IC 09) | | | | | | |
| 1 | < -0.01 | -1.25 | 0.219 | -0.16 | -4.70 | < 0.001 |
| Left Dorsal Attention Network (IC15) | | | | | | |
| 1 | < 0.01 | 0.33 | 0.746 | -0.34 | -5.77 | < 0.001 |
| 2 | 0.01 | 1.24 | 0.222 | -0.09 | -1.73 | 0.091 |
| 3 | -0.01 | -0.63 | 0.529 | -0.18 | -1.89 | 0.066 |
| 4 | 0.01 | 0.86 | 0.393 | -0.12 | -1.78 | 0.082 |
| 5 | < 0.01 | 0.77 | 0.445 | -0.23 | -4.20 | < 0.001 |
| 6 | 0.01 | -0.73 | 0.469 | -0.28 | -3.74 | 0.001 |
| 7 | 0.02 | 1.67 | 0.102 | -0.62 | -4.84 | < 0.001 |
| 8 | 0.01 | 1.59 | 0.119 | -0.24 | -4.06 | <0.001 |
| Auditory Network (IC 21) | | | | | | |
| 1 | < -0.01 | -0.33 | 0.74 | -0.73 | -5.21 | < 0.001 |
| Visual Occipital Network (IC 24) | | | | | | |
| 1 | < 0.01 | 0.08 | 0.93 | -0.14 | -3.71 | 0.001 |

**Table S1: No unique variance in rsFC explained by pain duration.** Model parameters of ranked-based regression models assessing the effect of pain duration and age on parameter estimates extracted form significant clusters. Grey color indicates values are not interpretable due to non-independence (Vu et al. 2016). IC, independent component.

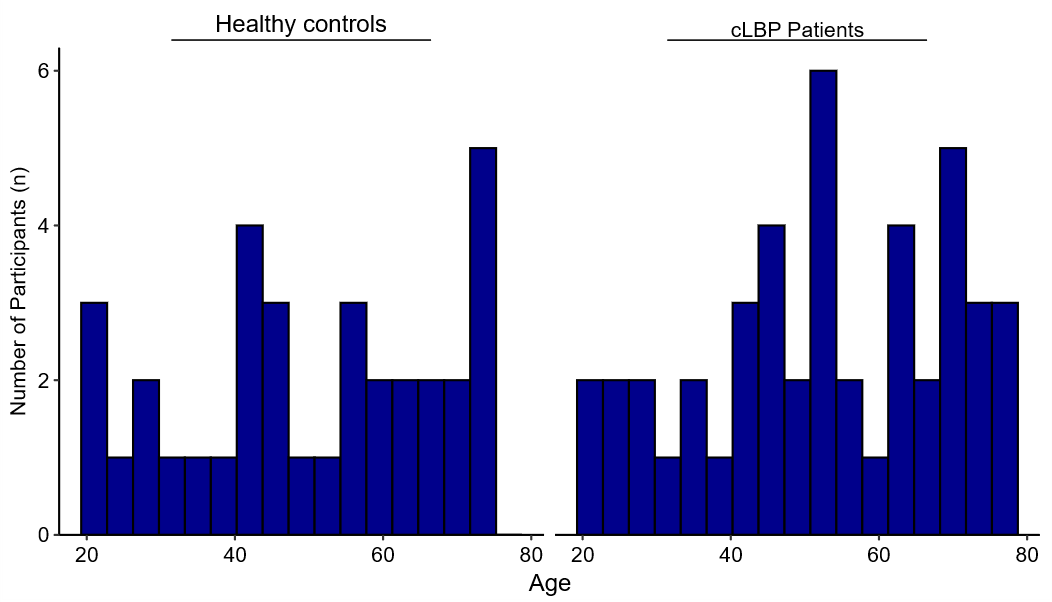

**Figure S1: Age distribution of cLBP patients and healthy controls.** Histograms of age for healthy controls and cLBP patients, with a bin width of 3.5 years. cLBP, chronic low back pain; HC, healthy controls.

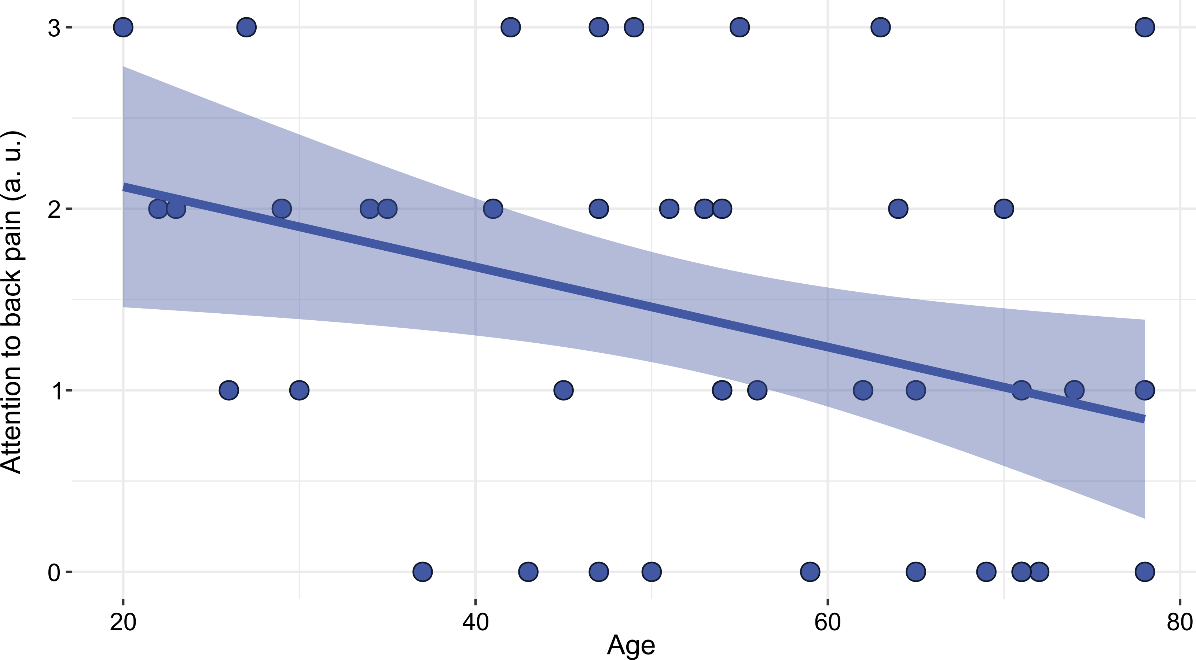

**Figure S2: Attention to back pain decreases with age.** Attention to back pain scores in relation to age for cLBP patients, with raw values (dots), the fitted regression line (solid line), and the 95% confidence interval (shaded area). A significant negative correlation with age in cLBP patients was observed (Kendall’s rank correlation: τ = −0.28, p = 0.016). Attention to back pain was assessed using a single-item questionnaire according to Akin-Akinyosoye et al., 2018, rated on a Likert scale ranging from 0 to 4. a.u., arbitrary units.
